# Mobile Lung Cancer Screening in Limited Resource Regions: The ProPulmão Project (BRELT3) Study Protocol

**DOI:** 10.1101/2024.07.25.24309976

**Authors:** Ricardo Sales dos Santos, Ricardo Figueiredo, Juliana P. Franceschini, César Augusto de Araújo Neto, Almério Machado, Bruno Hochhegger, Mario Claudio Ghefter, Ulisses Amancio Pereira Neto, Petrucio Abrantes Sarmento, Igor Barbosa Ribeiro, Daniel Augusto Xavier Carvalho, Felipe Passos, Caio Santos Holanda, Marcel Samuel Blech Hamaoui, Gustavo Borges da Silva Teles, Carolina Alves Neves, Helena Alves Costa Pereira, Jackline Pereira Leto, Adelmo de Souza Machado Neto, Audrey Cabral, Fernando Nunes Galvão de Oliveira, Clarissa Maria de Cerqueira Mathias, César Garcia Machado, Josiane Dantas Viana Barbosa, Marine Barbosa, Crislaine Gomes da Silva, Mariana Moreira de Silva, Lila Teixeira de Araújo, Álvaro A. Cruz

## Abstract

**Background:** Lung cancer is a highly aggressive disease that affected almost 2.2 million people worldwide and caused 1.8 million deaths in 2020. Smoking as well as being exposed to cancer-causing substances in the workplace are significant contributors to the chance of developing cancer. Developing countries encounter distinctive obstacles in the implementation of lung cancer screening because of extensive geographical and socioeconomic disparities.

**Objectives:** our primary objective is to describe tomographic findings in a high-risk lung cancer population in limited resource Brazilian areas. The secondary objectives are to quantify the frequency of pulmonary nodules (especially those falling into Lung-RADS categories 3 and 4) as well as the occurrence of lung cancer; to describe and analyze the challenges related to lung cancer screening programs in the context of Brazilian public health system; to explain the pulmonary function pattern and clinical characteristics of individuals diagnosed with moderate or severe emphysema by low-dose computed tomography (LDCT); and to evaluate the commitment of Community Health Assistants in actively recruiting the high-risk population.

**Methods:** This is a prospective cohort study. The study includes individuals in an age range of 50 to 80 years who are either current or former smokers and have a smoking history of at least 20 pack-years. They undergo LDCT with a planned follow up of 12 months. All adverse events are monitored and assisted. The classification of screening results is performed according to the Lung-RADS standards. Individuals classified in categories 3 and 4 receive additional diagnostic assessments and may have further testing.

**Expected results:** we expect this study shows the feasibility and effectiveness of lung cancer screening in people in situation of social vulnerability within limited resource settings, providing vital knowledge to reduce mortality and improve health outcomes. The project will generate relevant knowledge to inform policies on lung cancer screening within the Brazilian public health system, emphasizing the necessity of timely identification and action in limited resource settings.

**Outcome:** The study’s dissemination plan includes a website, social media, and participation in scientific conferences. Approval from the ethics committee has been obtained, and rigorous mechanisms are in place to guarantee the privacy of the data.

## Introduction

Lung cancer is an aggressive disease with a high global incidence, estimated at approximately 2.2 million new cases and 1.8 million deaths in 2020. [1] In the United States, tobacco product consumption is associated with 90% of diagnosed lung cancer cases. [2] Another significant factor is exposure to carcinogenic agents in occupational settings. In Brazil, it is estimated that 12,6% of the population are smokers. [3.4]

The substantial progress in lung cancer screening is attributed to the superior sensitivity and efficiency of Low-Dose Computed Tomography (LDCT) over conventional chest radiography. This advancement has resulted in decreased mortality rates and improved diagnostic accuracy, markedly enhancing the early detection and treatment of lung cancer. [6]. Additionally, several studies indicate that LDCT is cost-effective for lung cancer screening. [7–9]

The National Lung Screening Trial (NLST) in the U.S. involved 33 medical centers, comparing the effectiveness of annual LDCT screening to chest radiography in 53,454 high-risk individuals, demonstrating a 20% reduction in lung cancer mortality with LDCT.[8] The NELSON trial, conducted in the Netherlands and Belgium, involved 15,789 smokers or former smokers and found a 24% reduction in lung cancer mortality after 10 years of LDCT screening. [10]

There had been no lung cancer screening studies in Latin America until the Brazilian Early Lung Cancer Detection Trial (BRELT - ProPulmão) was initiated in Brazil. [11] The subsequent BRELT1 and BRELT2 studies contributed data and insights on lung cancer prevalence and screening efficacy. [12,13] The current BRELT3 study aims to investigate the challenges associated with implementing lung cancer screening programs in remote regions distant from major urban centers, where access to healthcare and resources is often scarce. Its goal is to broaden the reach and enhance the efficacy of screening among these underserved populations (Fig 1). Data resulting from this study will contribute to the characterization of the Brazilian population profile which is most exposed to lung cancer, given the heterogeneity and demographic dimensions of the country.

**Fig 1.** Lung cancer screening mobile unit, the “ProPulmão Respiratory Health Truck”. A. Prepared for transport; B. CT scan; C. technician performing examination; D and E. Prepared for patient care.

### Hypothesis

Implementing a mobile unit-based lung screening program in Brazil’s low resource settings is feasible and can significantly enhance lung cancer’s early detection, allowing for timely intervention.

### Objectives

The primary aim of this project is to describe tomographic findings in a mobile lung cancer screening program in a high-risk population in Brazilians’ remote regions. Secondary aims include report the prevalence of pulmonary nodules classified as Lung Imaging Reporting and Data System (Lung-RADS) categories 3 and 4 as well as the occurrence of lung cancer; investigate the challenges and barriers to lung screening in the public health system in developing countries; describe the lung function pattern and clinical features of patients with moderate or severe emphysema primarily identified through LDCT scans and explore the role and involvement of community health agents in engaging the local population in a lung screening program in developing countries.

## Methods

This is a prospective, open cohort study, with 1 year of follow-up, that includes participants with a history of smoking in inhabitants of selected cities in the state of Bahia that will host a mobile cancer screening unit. The start date of the recruitment period is August 08, 2023, and the duration of the recruitment period is one year. Candidates for the program will be identified through publicity in the local media and through an active search for possible candidates with screening criteria to be conducted by community health agents in the cities that will receive the mobile unit. The Brazilian Constitution guarantees universal, equitable, and comprehensive access to healthcare, highlighting the importance of understanding the challenges in facing this commitment within a nation of vast dimensions and significant social and demographic disparities. community health agents play a pivotal role within the Brazilian health system. They are tasked with assessing community needs, actively promoting health, and preventing disease [5]. Historically, community health agents have served as the crucial connection between healthcare professionals and the community. For this purpose, community health agents will undergo training to understand the fundamentals of lung cancer, the significance of screening for early detection, the main risk factors, and the guidelines for recommending screening.

In the study design, a non-probabilistic sampling method will be used, whereby all patients encountered by the research team will be approached and invited to participate in the investigation. The initial contact and complete interview of participants are carried out by a multi-professional team, conducted by regular nurses trained in the protocol, which is essential for minimizing costs related to population recruitment, avoiding unnecessary initial medical consultations.

One of the research team nurses will conduct the eligibility interview over the phone to identify and invite the potential participants. Three thousand individuals will be consecutively enrolled. The study will include those who meet the following eligibility criteria: aged between 50 and 80 years, self-reporting as asymptomatic, currently active smokers with a history of at least 20 pack-years, and former smokers who have ceased smoking within the last 15 years. The exclusion criteria comprehend:

- Individuals unable to perform the LDCT examination;
- Intolerance to the horizontal supine position for more than 10 minutes;
- Presentation of highly suggestive symptoms of lung cancer;
- Diagnosis of serious heart disease on multiple medications;
- Diagnosis of severe lung disease, on multiple medications and/or requiring home oxygen therapy;
- History of chest radiotherapy;
- Pregnancy.

The included people will participate in the initial program interview conducted by one of the program’s nurses. This interview will include a detailed medical history to assess personal and family history, exposure to major risk factors for chronic respiratory disease, smoking history, and symptoms. At the end of the interview, following free and informed consent form signatures, participants will be sent directly to the LDCT without a pre-examination medical consultation.

The outcome variables include the results of the first LDCT, performed after the participant inclusion in the study, and, in specific cases, new LDCT performed after 3 or 6 months, depending on the characteristics of the findings and the results of the spirometry, that will be performed on participants who present emphysematous findings in LDCT classified as moderate or severe.

The multidisciplinary team will monitor patients over a period of 12 months. Visits include the following procedures: LDCT, spirometry (for participants with pulmonary emphysema data classified as moderate or severe on LDCT), and medical consultations. Details of each step are described in Fig 2.

**Fig 2.**
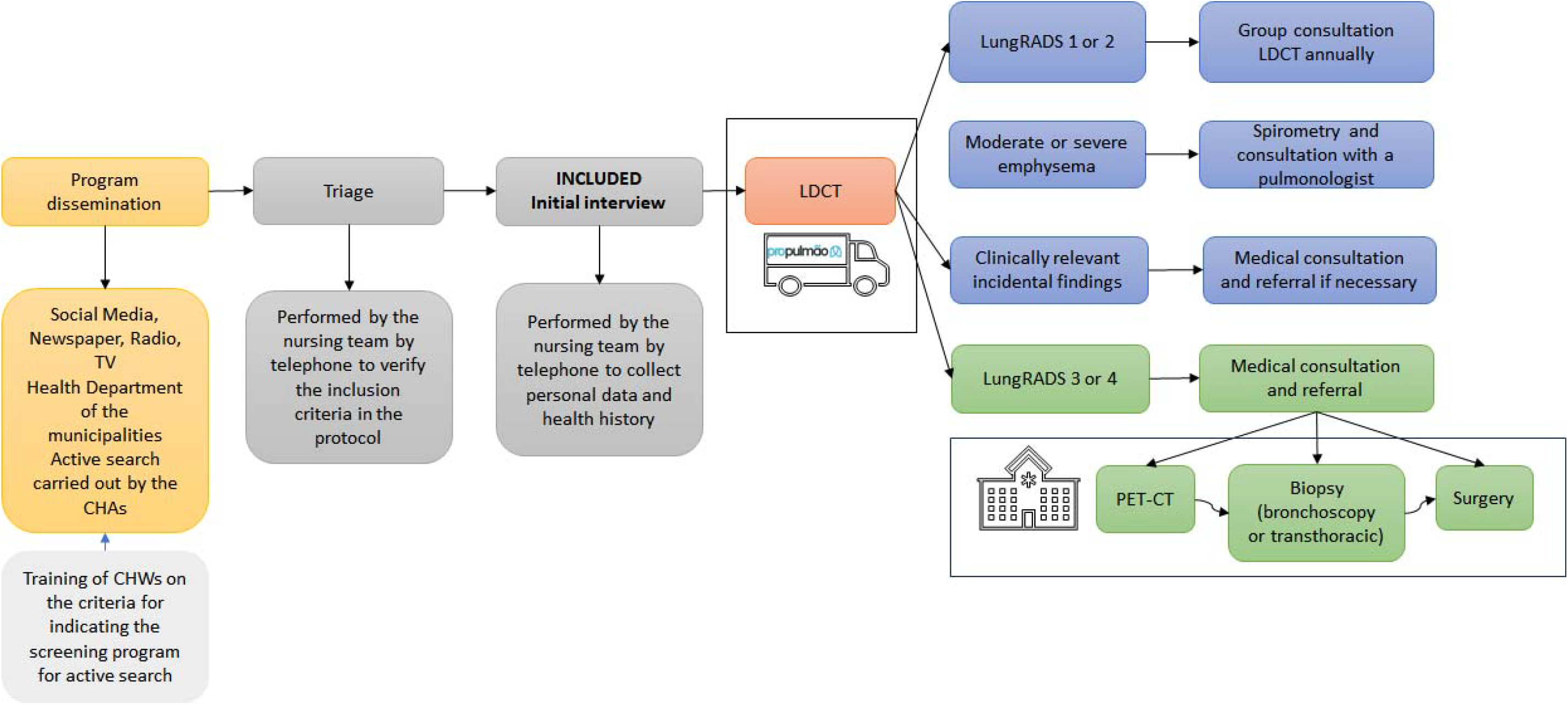
Lung cancer screening mobile unit workflow based on LungRADS classification.

All LDCT exams will have LungRADS classification. LDCT results will be provided to participants in a group session orientation for all LDCT classified as LungRADS 1 or 2. Examinations classified as LungRADS 3 or 4 are discussed in an online tumor board meeting (Screening Board Committee) by a specialized medical team of radiologists, pulmonologists, thoracic surgeons, and oncologists.

To facilitate case analysis and discussion, clinical data and images are previously sent to the Screening Board Committee (SBC) and presented virtually during the online meeting. Following the presentation, SBC participants can vote using an online form with the main lung nodule approaches listed. When the suggested nodule management receive 70% or more of the SBC votes, the specific approach is considered to the participant. In cases of disagreement, the case is discussed further by the multidisciplinary team to determine the most appropriate management.

Based on this discussion, further testing may be recommended following international recommendations to diagnose or rule out the possibility of lung cancer. Follow-up options are based on recommendations from the NCCN Lung Cancer Screening Guidelines and the American Society of Radiology according to the Lung-RADS staging categories. According to these recommendations, categories 1 and 2 include nodules with a benign appearance for which annual screening should be continued. LungRADS 3 or 4 should be evaluated based on size, solid component, and growth rate, and repeat imaging in three to six months or consider positron emission tomography (PET-CT) for nodules with a solid component greater than 8 mm. In higher suspicion cases, recommendations may include transthoracic puncture, bronchoscopy, or surgery for histopathologic confirmation of the possible diagnosis of lung cancer. Final lung nodule management will be defined by the medical practitioner in a share decision making process with the patient, adding local or social aspects not included on the initial SBC analysis.

The program will maintain periodic contact with participants to ensure adequate follow-up over time. The follow-up period will be essential to monitor the development of nodules, perform additional tests as necessary, and ensure continuity of care.

During the follow-up period, safety-related variables will be evaluated, emphasizing monitoring serious adverse events or complications, classified in causality and intensity according to the Naranjo and WHO causality algorithm.

Additionally, secondary variables will include efficacy measures related to expected study outcomes. When evaluating efficacy, the importance of the positive predictive value of the LDCT method is highlighted. At least 6.3% of the participants assessed will likely have Lung RADS 4 categories and between 0.7% and 3.1% of lung cancer at the end of follow-up.

### Quality control

LDCT exams will be evaluated by senior radiologists trained in the LungRADS classification, and trained staff will perform spirometry. The anamnesis and selection of candidates are carried out using the same criteria established in standardized medical consultations for screening.

For data quality control, a data bank using RedCap platform was performed, where Case Report Forms will be fulfilled with demographic, sensible and exams data from all participants and this information will be stored and monitored in the mentioned bank and presented in real time to the research team through a Power BI dashboard.

Often the imputed data are checked by sampling and in case of errors identification, the data entry team receives retraining as needed.

### Sample size

Initially, we plan for this study to include 3000 participants to evaluate our primary outcome based on convenience sampling.

Based on the population pyramid projection and the prevalence of smoking in the Northeast region of Brazil, it is anticipated that a significant portion of individuals in the communities visited by the mobile unit will be at risk. Assuming an average population of one hundred thousand inhabitants for these variables and considering that approximately 19% of the population is over 50 years old, we can estimate a total of 19 thousand individuals within this age group.

Considering the current prevalence of smoking in Bahia, which stands at about 7%, and factoring in an additional 10% representing probable ex-smokers, we arrive at an estimated population at risk of nearly 3 thousand individuals for each community visited. It’s important to note that this estimate excludes patients already diagnosed with chronic diseases and undergoing regular treatment.

### Statistical analysis

We intend to use Statistical Package for the Social Sciences (SPSS) version 29 to perform statistical analysis. Descriptive statistics will be used to summarize the demographic characteristics of study participants. Categorical variables will be summarized in absolute and relative frequencies (percentages). Information on numerical variables will be expressed as means, standard deviations (SD), medians, and minimum and maximum values.

The following statistical methods will be used to analyze the results: Chi-square test, Mann-Whitney test, Student’s t-test, and ANOVA with Bonferroni’s supplementary test, among others. For all statistical tests, the significance level adopted will be Type I error <0.05 or 5%.

### Ethics

This research strictly follows the Brazilian Ethical guidelines (Resol 466/12; RDC 837/2023) and international guidelines, such as ICH E6 (R2), and Good Clinical Practices-Document of Americas), ensuring scientific integrity, reproducibility, traceability and reliability of the study. Participants are informed about the objectives and procedures, and written informed consent is obtained. Ethical measures guarantee data privacy and confidentiality, respecting fundamental scientific research principles and the Brazilian Data Protection Law (LGPD 13709/2018). This study is designed to adhere to the highest standards of human research practices, prioritizing the transparent disclosure of all outcomes [14]. The commitment to good practices seeks transparency in the disclosure of results, regardless of their nature. This approach aims to enhance both the credibility and the replicability of the study’s findings. The study is approved by the SENAI-CIMATEC Institutional Review Board through a CAAE number 67431523.6.0000.9287, with a financial support from the Bristol-Myers Squibb Foundation for the development of the mobile unit and LDCTs, Boehringer Ingelheim Brazil for spirometry, Diagnósticos da América S.A. (DASA) for CT reports, AstraZeneca Brazil & Lung Ambition Alliance for educational activities and Ethicon Brazil and Panther Brazil for medical devices.

### Dissemination plan

The BRELT3 will implement a comprehensive communication strategy to actively disseminate the study findings. The communication strategy of this study in the public health system, in constant development with the research team, involves a wide-ranging dissemination plan that includes the creation of the ProPulmão website for information sharing, the use of social media platforms for engagement, participation in scientific conferences to connect with the research community, and conducting technical workshop tours. Furthermore, the strategy incorporates issuing press releases to reach a broader audience, hosting a significant high-level event to discuss key findings, and organizing workshops and roundtable discussions facilitated by a professional communication agency to foster dialogue with the scientific community and disseminate knowledge. In addition, the research team will develop meetings in the cities where participants were included, with the aim of informing the population and health government managers about the study findings and also raise awareness, as well as motivation to lung screening public policies.

### Risks

Radiation exposure from LDCT screening and subsequent diagnostic procedures contributes to increased cumulative radiation. Nonetheless, the individual risk of developing lung cancer due to radiation in screening programs is relatively minor (0.2%-1%) when compared to the risks associated with smoking (approximately 16%). [15]

Patients who undergo invasive diagnostic procedures following lung cancer screening with LDCT in real-world settings may experience complications. The majority of these complications are classified as minor or intermediate in severity.

The data from the medical record and interview will be confidential and restricted. Access to the study database will be limited to principal investigators. Although best efforts will be mobilized to ensure the anonymity of all records, there is a potential risk of accidental breach of anonymity.

## Strengths and limitations

To our knowledge, this will be the largest study to assess the prevalence of lung nodules in this type of at-risk population with low access to early diagnosis.

The criteria for inclusion in lung cancer screening programs are subject to ongoing revision. Registers of individuals who are currently smoking and not eligible under the existing USPSTF (U.S. Preventive Services Task Force) guidelines may facilitate subsequent research involving this demographic.

Individuals presenting with significant symptoms are directed to local health services since the focus of the current study is on preventive measures. Consequently, evaluating and managing their present symptoms should be addressed within the standard healthcare system. Identifying symptomatic cases could represent a more significant overload on the local health system. A potential limitation of the current screening implementation study is its limited capacity to absorb potential cases of advanced-stage lung cancer.

## Impact

Lung cancer, the most lethal form of cancer globally, is an escalating concern in Latin America. The increasing incidence of this disease poses numerous challenges to the region’s economies, particularly regarding constrained resources to address the healthcare demands in developing countries.[16] Implementing a lung cancer screening program in low-to-moderate-income countries presents significant barriers. This study aims to provide insights into the feasibility and effectiveness of screening within underserved populations in remote regions distant from major urban centers in developing countries.

## Data Availability

No datasets were generated or analysed during the current study. All relevant data from this study will be made available upon study completion.

